# The father’s singing voice may impact premature infants’ brain more than their mother’s: A study protocol and preliminary data on a singing and EEG randomized controlled trial (RCT) based on the fundamental frequency of voice and kinship parameters

**DOI:** 10.1101/2024.09.29.24314570

**Authors:** Efthymios Papatzikis, Kyriakos Dimitropoulos, Kassandra Tataropoulou, Maria Kyrtsoudi, Elena Pasoudi, John M. O’Toole, Angeliki Nika

## Abstract

This article presents the study protocol for a randomized controlled trial (RCT) investigating the impact of singing on the brain activity of premature infants in the Neonatal Intensive Care Unit (NICU). The study focuses on how the differentiation of voices, as defined by the fundamental frequency (F0) shaped by biological sex and kinship, influences neurophysiological responses when measured by electroencephalography (EEG). Premature infants, who are highly sensitive to auditory stimuli, may benefit from music-based interventions; however, there is limited understanding of how voice variations between male and female caregivers, and whether they are biologically related, affect brain activity. Our protocol outlines a structured intervention where infants are exposed to singing by four facilitators - a male music therapist, a female music therapist, the mother, and the father - and includes two singing stages: a sustained note (A at 440 Hz) and a 90-second lullaby, both interspersed with silent periods to allow for baseline measurements. EEG recordings track brain activity throughout these sessions, followed by quantitative EEG (qEEG) analysis and thorough statistical computations (e.g., mixed-effects models, spectral power analysis, and post-hoc tests) to explore how these auditory stimuli influence brain function. Preliminary data from five infants show that maternal singing elicits the highest delta spectral power in all measured conditions except during the ‘lullaby song’, where paternal singing elicits the highest effects followed by the male music therapist and then the mother. These early findings highlight the potential influence of parental voices, particularly the fathers’ voice, on neonatal brain development, while the detailed study protocol ensures rigor and replicability, providing a robust framework for future research. Additionally, this protocol lays the groundwork for exploring the long-term effects of music-based interventions, with the goal of improving neurodevelopmental outcomes in premature infants through tailored auditory stimulation. (clinincaltrials.gov unique identifier: NCT06398912)

## Introduction

In the evolving landscape of neonatal neurology, the focus has increasingly shifted towards understanding the complex interplay between the environment and the developmental outcomes of neonates, especially those in critical care settings (Blackburn, 1998; Santos, Pearce, & Stroustrup, 2015; Bonkowsky, deVeber, Kosofsky, & Child Neurology Society Research Committee, 2020). Globally, a significant number of newborns (just over 13 million) are born prematurely, and many of these vulnerable infants spend their early, formative days in Neonatal Intensive Care Units (NICUs) (World Health Organization, 2022). Advances in neonatal medicine have dramatically improved the survival rates of these infants (e.g., Qattea et al., 2022; Gebreheat & Teame, 2022) yet challenges persist in ensuring their long-term neurological health and development.

Structured sound interventions, approached as non-pharmacological supplementary treatments, have shown promise in improving health outcomes for this vulnerable population in the context of the NICU. For example, white noise has been found to mimic the intrauterine environment, aiding in the stabilization of sleep patterns and reducing stress responses in premature infants (Akiyama et al., 2021). The use of heartbeat sounds has also been instrumental in providing a rhythmic and familiar auditory stimulus to these infants, facilitating physiological regulation and promoting a sense of security (Zhang & He, 2023; Alemdar & Özdemir, 2017). Similarly, environmental sounds, such as gentle rain or soft wind, have been shown to create a soothing and more natural auditory environment, contributing to the reduction of the clinical and mechanical noise of the NICU (Iyendo, 2017; Praskach, 2009; Almadhoob & Ohlsson, 2020). Finally, passive listening to more organized sounds or music has been widely documented to enhance infants’ physiological and emotional well-being (Papatzikis et al., 2024a), with studies demonstrating the positive effects of music on stress reduction, pain management, and weight gain (Pölkki & Korhonen, 2012; Caine, 1991; Arnon et al., 2006; Loewy et al., 2013).

### Delivering (live) music interventions in the NICU

While NICUs are pivotal for medical care, they also introduce infants, especially the premature ones, to unique sensory environments (Reuter et al., 2023; Brown, 2009; Vicencio et al., 2023). These environments differ significantly from the acoustical environment of the mother’s womb or the typical domestic environment, and can profoundly influence developmental processes (Kvaratskhelia et al., 2023; Chung et al., 2020; Cook et al., 2023). Therefore, the prescription and implementation of music in NICUs should adhere to tailored and sensitive intervention protocols based on well-studied (Papatzikis et al., 2023) and biology-based facts (Papatzikis, 2024) to avoid disrupting the developmental trajectory of the infant and potentially act as a developmental catalyst.

For instance, regarding the administration of any sound stimulus in the NICU context, careful moderation is expected to prevent overstimulation and ensure the maintenance of physiological stability in neonates. The extreme sensitivity of premature infants to auditory stimuli necessitates that all sound related stimuli in the NICU should adhere to certain acoustical regulations as defined by specific regulatory medical bodies or global standards (White et al., 2013; EFCNI, Sizun, Hallberg, et al., 2018). These regulations are designed not only to prevent adverse fluctuations in vital signs such as heart rate and respiration – ultimately resulting in stress responses in the infant – but also to optimize the therapeutic benefits of environmental and intentionally administered sound stimuli. Additionally, they aim to minimize potential disruptions to proper biomechanical development, such as damage to the hair cells in the infant’s inner ear. These specialized ear cells are very sensitive to apoptosis events and can take place due to excessive sound stimulation (Hu et al., 2000; Kamio, Watanabe, & Okubo, 2012; Gerhardt & Abrams, 2000).

Furthermore, the timing of sound-related interventions should be strategically planned not only to align with each infant’s individual feeding and sleep-wake cycles (Bueno & Menna-Barreto, 2016) – as these factors can potentially interfere with the beneficial effects of musical interventions – but also to ensure they occur during quieter periods in the NICU, allowing for safe and effective delivery (Almadhoob & Ohlsson, 2020). In particular, live singing interventions must be carefully calibrated and monitored to stay within safe sound limits set by medical regulations (see EFCNI, Sizun, Hallberg, et al., 2018). These regulations generally stipulate that sound levels outside the incubator should not exceed a continuous average of 45-50 dB, with transient peaks not surpassing 65 dB. Unfortunately, even highly trained professional singers, when attempting to maintain the lowest possible sound pressure levels (SPLs), may still exceed these limits. A study by Akerlund & Gramming (1994) found that trained female singers could only produce minimum SPLs around 51.5 dB at low pitches, which already slightly exceeds the upper recommended continuous sound level in NICUs.

Finally, a seamless integration of any musical intervention into the infants’ overall care regimen requires close coordination with the medical team, as this collaborative effort is vital for aligning the intervention sessions with the already established medical protocols and tailoring them to meet the specific health requirements and medical conditions of each infant (Kuo et al., 2018). Such integration helps to ensure that the music intervention sessions are not only safe but also effectively support the broader therapeutic goals set forth by the healthcare providers, which especially in this sensitive and demanding in terms of physical development context require the parental involvement as another critical aspect according to the latest clinical research (Dokkum, Fagan, Cullen, & Loewy, 2023; Haslbeck & Bassler, 2020).

### The role of Kinship in the NICU Singing process

Prematurity poses significant risks to child development on social, behavioral, and cognitive levels (Guidry-Jorgensen et al., 2011). Most importantly, the early bond between the biologically related caregiver and the infant is crucial for the latter’s cognitive development, influencing the child’s self-representation and future interpersonal relationships (Borghini et al., 2006; Vrticka & Vuilleumier, 2012). However, in the NICU, the typical biological processes of feeding, caring, and bonding are often disrupted due to the infant’s medical complexities, leading to stress for both infants and parents (Shoemark et al., 2015).

Infant attachment issues are prevalent among parents who have undergone significant trauma, impairing their sensitivity towards their baby and heightening their vigilance about the baby’s condition (Coppola et al., 2007; Shoemark et al., 2015). Research indicates that premature infants often exhibit more passive behavior, less positive affect, and reduced facial expressions during their interactions with their biological caregivers (Bozzette, 2007). Correspondingly, parents of premature infants tend to be less sensitive in these interactions, negatively impacting the infant’s developmental trajectory (Field, 1995; Forcada-Guex et al., 2006; Muller-Nix et al., 2004). Conversely, sensitive caregiver attitudes and parent-infant synchronization during interactions have been shown to be protective and beneficial for the development of premature infants (Forcada-Guex et al., 2006; Newnham et al., 2009; Treyvaud et al., 2012).

In this context, singing can serve as a parent-infant synchronization intervention, fostering emotional bonds and providing comfort to both infants and their parents (Lense, Shultz, Astésano, & Jones, 2022; Fancourt & Perkins, 2018). Additionally, singing in the NICU can stabilize an infant’s physiological state, including heart rate and breathing, and promote better sleep patterns (Kobus et al., 2021; Yakobson et al., 2021). It can also serve as a conduit for emotional expression and connection, offering a sense of normalcy and control for parents who might otherwise feel helpless in the highly medicalized environment of the NICU. As a result, through singing, parents can engage in meaningful interaction with their infants, reinforcing their role as primary caregivers and fostering a secure attachment (Shoemark et al., 2015).

However, parents are not the only caregivers in the NICU, and for this reason they are not the only ones who may be able to provide singing sessions. Multiple research studies have shown that caregivers with no immediate kinship – like nurses, physicians and other clinical staff – can also effectively use singing to support developmental and medical outcomes (Liu et al., 2023). This dynamic is, of course, complex and influenced by factors such as the differentiation of kinship and the recognizability of the caregiver’s voice. The fundamental frequency of their voice, which differs from that of the mother and father, may be less familiar to the infant (Spence & Freeman, 1996; Mills & Melhuish, 1974; Mai et al., 2012).

### The Fundamental Frequency of Singing

In the context of the NICU, understanding the technical features of the singing voice, including its fundamental frequency (F0) as opposed to pitch, becomes increasingly vital, not only due to the auditory sensitivity of premature infants but also because of the developing nature of their central nervous system, which is highly susceptible to the variability of external stimuli as they grow older.

The fundamental frequency (measured in Hertz) refers to a physical property of sound waves – specifically, the number of vibrations per second produced, for example, by the violin string or, in our case, the vocal cords (Hirst & De Looze, 2021; Gerhard, 2003). Pitch, on the other hand, is the perceptual correlate of fundamental frequency; it describes how the brain interprets and perceives the fundamental frequency (Hirst & De Looze, 2021). While F0 is an objective measurement, pitch is a more subjective experience; however, for most periodic sounds, the fundamental frequency closely corresponds to the perceived pitch (Hirst & De Looze, 2021; Gerhard, 2003). As a result, F0 is often used as a proxy for pitch in acoustic analysis and research.

Given the critical role of fundamental frequency (F0) in shaping the auditory environment for infants, particularly in a singing context, it is important to understand the differences in F0 between male and female voices. These differences, driven by anatomical factors such as the length of the vocal tract and the size of the laryngeal cavities, result in male voices typically having a lower F0 – around 100-120 Hz – compared to the higher F0 of female voices, generally in the 200-220 Hz range (Titze, 1989; Coleman, 1976; Fant, 1966). Such distinctions may influence how infants respond to structured sound stimuli, especially in light of the well-documented human sensitivity to voice discrimination based on biological sex (Latinus & Taylor, 2012). The fundamental frequency serves as the foundation upon which harmonic structures are built, guiding the singing experience and creating a scaffold for the infant’s brain to process the embedded complex auditory information. These differences in F0, therefore, may not only influence how infants respond to the singing environment in the NICU but may also convey biological and kinship-specific cues that can affect neural functioning and development in this sensitive population.

### Study Objectives

The F0 of singing and its impact on neonatal brain development remains under-explored, particularly in the context of NICU environments. Current literature lacks detailed insights into how F0 variations, influenced by the biological sex and kinship of the singing source, affect the short-term neurophysiological responses of premature infants. Therefore, this study, aiming to address this research gap, is set to investigate the specific mechanisms through which different F0 profiles from a combination of human voices and kinship dynamics (i.e., mother, father, male, and female music therapists) influence the brain activity of NICU infants.

The primary research questions of this study are: *Does a structured sound and music-based intervention (mentioned as musical intervention from now on) from the two different biological sex profiles (i.e., male, female) based on their F0 result in a differentiated short-term electroencephalographic effect footprint in NICU infants? Furthermore, is there differentiation if their kinship (mother, father) with the newborn becomes a factor?*

Our research hypotheses are as follows:

- *H1*: We hypothesize that when premature newborns are exposed to a musical intervention provided by their mother (high fundamental frequency – biological kinship) compared to the father (low fundamental frequency – biological kinship) or an unrelated male/female music therapist, they show statistically more visible short-term positive oscillatory differentiations in their brain activity.
- *H0*: We assume that there is no interaction between the fundamental frequency and the human profile (mother, father, man, woman) in the infant’s reactions, and that neither the fundamental frequency nor the human profile has a primary effect.

To answer the research questions and study its hypotheses, this study will utilize electroencephalography (EEG) to monitor and analyze the brain activity of NICU infants. By comparing EEG patterns across different music delivery conditions of F0 and kinship source (i.e., music facilitators), the findings are expected to provide valuable insights that can be leveraged to enhance neurodevelopmental outcomes in premature infants.

## Methods

### Study design

Between 06.2023 and 12.2025, all premature newborns who meet the specific inclusion criteria (detailed below) in the NICU of the ‘Panagiotis and Aglaia Kyriakou’ Children’s Hospital in Greece will be screened, and those who qualify will be invited to randomly participate in either the intervention or control group of our study. During their stay in the NICU, the music intervention group will be exposed to a structured musical intervention – including varying investigative conditions of silence and singing – for four days within one week. Meanwhile, the research team will ensure that the control group is not exposed to any musical stimulation during the same period. Randomization and allocation of the eligible participants to one of the two groups (control or intervention) will be performed using a stratified randomization technique to account for parental availability (i.e., not all parents are living in the region the intervention is taking place, or parental working schedules do not fit towards an intervention scheduling). For this reason, participants will be stratified into two groups based on whether their parents are available to participate. Within the ‘available parents’ stratum, participants will be randomly allocated to either the control or intervention group (by a lotarry), ensuring a balanced distribution of parental availability across both groups. For the ‘non-available parents’ stratum, participants will be assigned only to the control group, as this group involves no active parental participation. This stratified approach ensures that the study properly tests the influence of parental involvement in the intervention group, while the control group includes infants regardless of parental availability. We believe that this randomization approach maintains the integrity of the study by ensuring a balanced group assignment, while also addressing practical and ethical considerations of the specific NICU setting. By the same token, blinding of the intervention is also difficult to implement in this research context. In terms of measurements, electroencephalography (EEG) will be used to record the newborns’ brain activity throughout their participation in the research study (both intervention and control group), while video data will be also recorded, forming an auxiliary part to the EEG data collection process. The schedule of enrolment, intervention and assessment can be seen in Figure 1 while a flowchart of the study design is shown in Figure 2.

**Figure 1.**
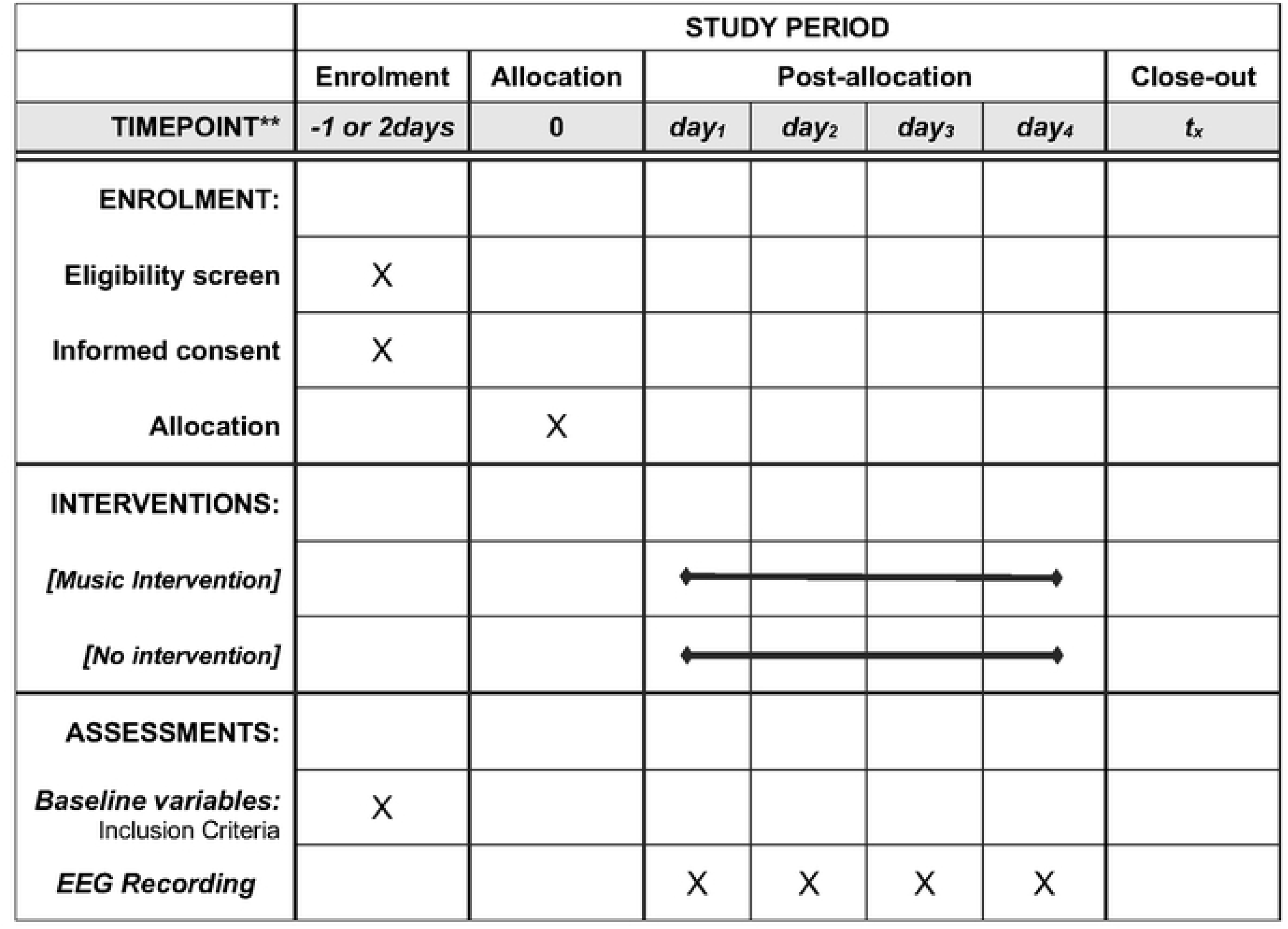
Schedule of enrolment, intervention and assessment

**Figure 2.**
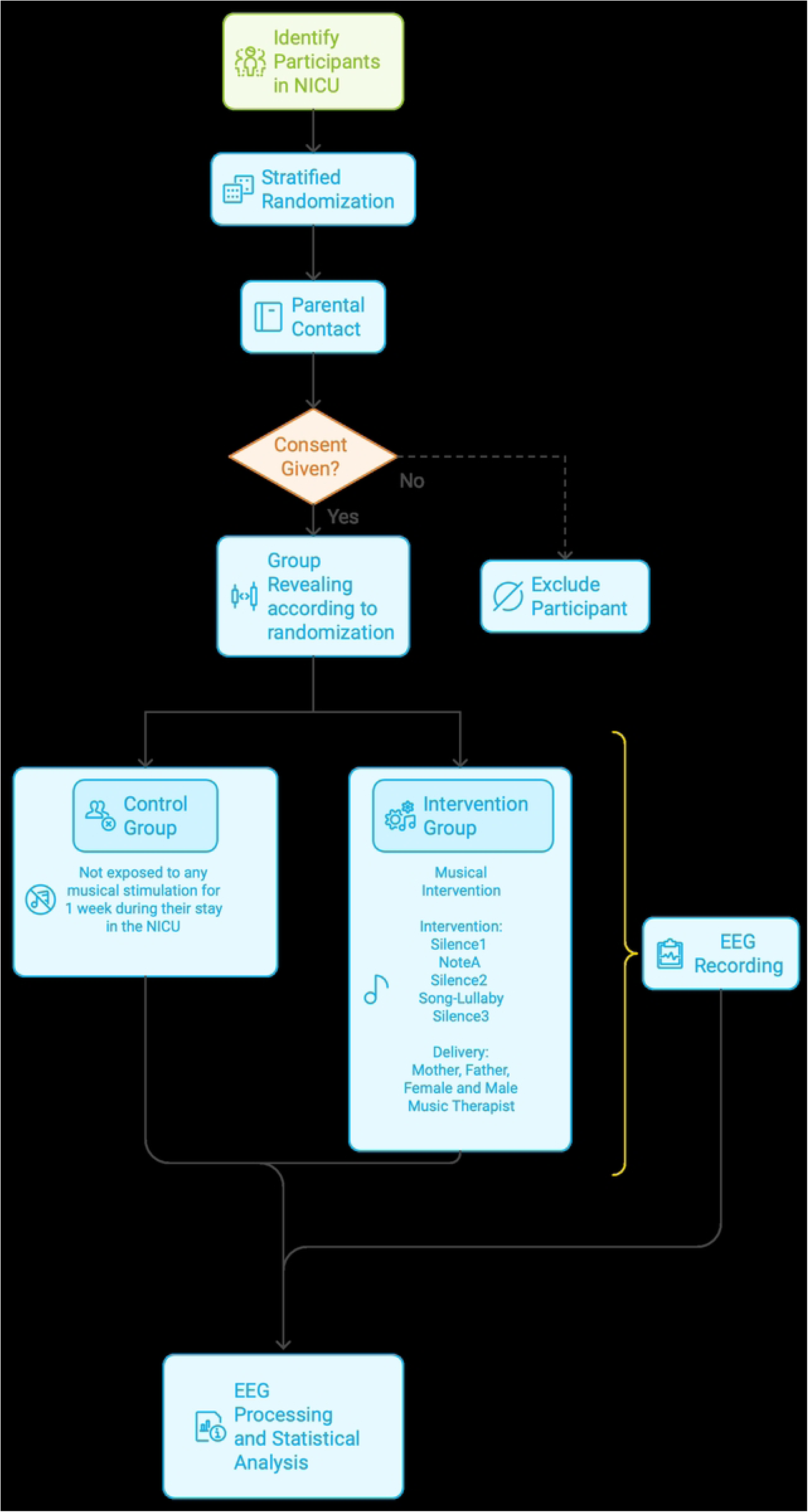
Study Protocol Flow Chart

In this design, although the EEG will be the primary tool of data collection and analysis, the rationale for incorporating video data is multifaceted. Firstly, the video recordings will complement the electroencephalogram (EEG) recordings in offering a comprehensive parallel view of the infants’ neural and behavioral responses. Furthermore, the videos will serve as a tool for quality control by ensuring that the prescribed research protocol is uniformly followed by all facilitators, contributing to the scientific integrity of the research. Finally, the videos will allow for a later examination of any synchrony or coordinated interactions between the infant and the facilitator, shedding light on phenotypical – not able to be measured by the EEG – aspects of the intervention.

The musical intervention will be delivered by four different profiles of individuals-facilitators: a male music therapist, a female music therapist, the mother, and the father of the newborn. During the delivery of the intervention (explained in more detail below), there will be five ‘conditions’ measured: Silence1 (180 seconds), Note A (60 seconds), Silence2 (180 seconds), Song (90 seconds) and Silence3 (180 seconds).

It is important to note that the study protocol will be conducted in a dedicated room, largely isolated from the ambient noise typically found in the rest of the NICU. This aspect of the study design is crucial for two main reasons:

Firstly, it will ensure the comfort and safety of the participating newborns. All actions taken in the context of the NICU require adherence to the recommended standards for the NICU sound environment. Therefore, the importance of a well-managed acoustic environment in reducing stress and discomfort for our participating infants is of paramount importance. Such a consideration has been consistently highlighted both for clinical and research settings alike (European Standards of Care for Newborn Health; EFCNI, Sizun, J., Hallberg, B., et al., 2018) and a sound-controlled room within the NICU seems to be the only viable option for achieving the necessary level of sound management for our participants, without disrupting other clinical operations. More specifically, in the NICU context, when considering the operational sound as outlined by White, Smith, Shepley, et al. (2013), the combination of continuous background sound and operational sound should not exceed an hourly L_eq_ of 45 dB and an hourly L_10_ of 50 dB, both A-weighted slow response, with transient sounds or L_max_ not exceeding 65 dB. Our study, with sound exposure lasting less than 3 minutes per hour, will strictly adhere to these guidelines. However, the transient nature of singing, distinct from the continuous and better-controlled sound pressure levels typically found in continuous speech, might lead to brief instances where we slightly exceed these thresholds by about 15 dB. According to evidence, even professional singers, and more so untrained individuals like the parents participating in our study, are not able to produce transient peak Sound Pressure Levels (SPLs) below 78-79 dB when singing softly (Boren, Roginska & Gill, 2013). Therefore, we believe that a dedicated room will significantly aid in ensuring the correct application and viability of the aforementioned recommendations – as compared to the open NICU floor – without pushing to the maximum the soundscape limits.

Secondly, recent research, including a study by Restin et al. (2021), indicates that incubators can be significant sources of noise, especially in the open environment of the NICU floor. To mitigate this issue, we plan to use an open cot/radiant warmer (as depicted in Figure 3) in the dedicated room when applicable, or alternatively, bring the entire incubator into the room. This approach will help minimize or eliminate noise from the incubator that might reach the infant. As an additional precaution to all the above, a dB measuring device will be also stationed in the dedicated room, outside the incubator where applicable, to monitor noise levels throughout the intervention.

**Figure 3.**
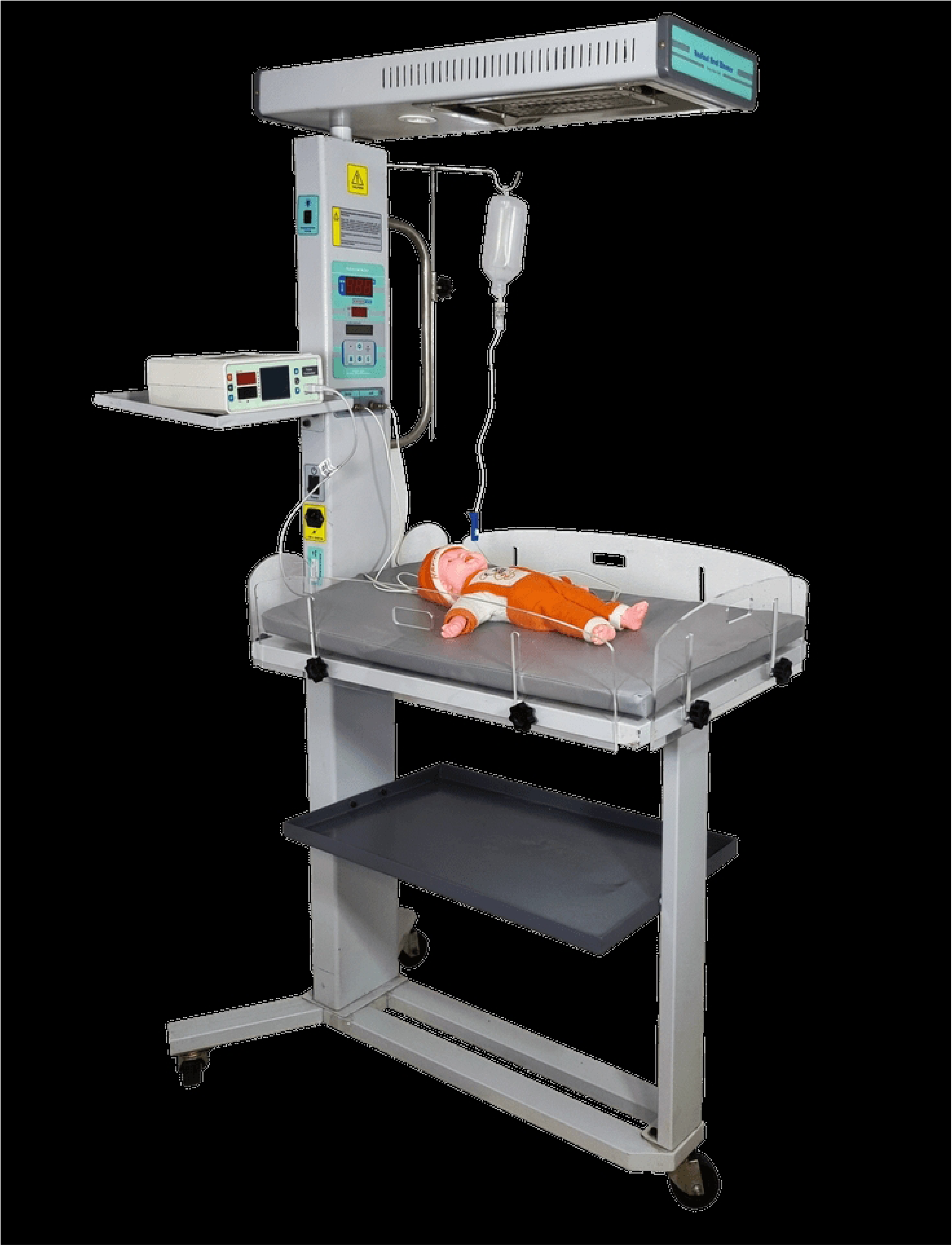
Open cot – Radiant warmer

#### EEG Tool, Data Collection and Processing

EEG is an electrophysiological imaging tool commonly used in medicine and research. The electroencephalogram measures changes in electrical dynamics resulting from the formation of electrical dipoles during nerve stimulation. The EEG signal consists of several brain waves that reflect electrical activity in the brain, based on electrode placement and function in adjacent brain areas. EEG-based changes in response to interventions have become a popular method for investigating brain function in patients with various brain and mental health disorders, as well as in healthy individuals, while the last few years, this tool has been successfully applied to premature newborns and full-term infants, too (Wu J et al., 2013).

In our study, we decided to use the **nëo™** EEG monitoring unit by ANT, a specialized tool designed for both routine and advanced neonatal care in NICUs. Eight high-precision channels found on a clinical-grade infant-sized cap are applied on our participants, connected to a research-grade amplifier with an input impedance exceeding 1 GΩ and advanced noise reduction. We chose this EEG system as it supports neuroprotective intervention screening, offering a maximum sampling rate of 512 Hz and 24-bit resolution.

For the raw EEG data preprocessing part, we have configured a pipeline specialized for the premature infants (as shown in supplementary file SF1, and presented in Papatzikis et al., 2024b; Papatzikis et al., 2024c; Papatzikis et al., 2024d), while for the processing part we followed and adapted to our protocol a specialized for the premature infants’ population quantitative EEG (qEEG) software developed for neonatal EEG by O’Toole & Boylan (2017). This software has been selected as our preferred EEG processing tool due to its highly informative, detailed, and objective output. Coupled with our preprocessing pipeline, this tool proved to be particularly advantageous for our research, as it allowed for the analysis of various EEG features such as spectral power, coherence, and connectivity among EEG channels, taking into consideration the intervention’s musical conditions and their short delivery profile. We aimed for this comprehensive EEG setup to enhance our ability to interpret the complex EEG data from our participants in greater detail, as these infants, compared to term infants, toddlers, and adults, exhibit unstable physiological and electroencephalographic states – such as signal continuity/discontinuity (Pavlidis et al., 2017; Pavlidis et al., 2020) and increased artifact generation (Wallois et al., 2021; Britton et al., 2016) – making it significantly more challenging to identify patterns and trends in the EEG data collected following our musical protocol.

#### Participants

Premature newborns who meet the inclusion criteria outlined below are eligible for participation in our study. These criteria have been carefully selected to ensure that the infants included have the appropriate health profile for safe participation and that the study results will be as accurate and relevant as possible. More specifically, to be eligible, newborns must have been born at 32 weeks of gestation or less and weigh at least 1000 grams, while not exceeding 38 weeks of postmenstrual age (PMA) during the intervention and measurement period. It is essential that potential participants do not have auditory insufficiency in the 8th cranial nerve or excessive pathology in the brainstem, while they must also have negative bilateral ‘transient evoked otoacoustic emissions’ – both verified during the postnatal screening by the in-house audiologist.

The eligible participants must also be in stable health, with no imminent risk of death, and must not be sedated or require sedative drug administration during the study. Any congenital, genetic, or chromosomal abnormalities, neonatal sepsis, or nuclear jaundice will also lead to exclusion, as these conditions could potentially influence brain function and interfere with the study’s results. Additionally, the participants must not be exposed to maternal use of illicit drugs during pregnancy, while mechanical ventilation used for their life support – that generates excessive noise, such as high-frequency oscillatory ventilation – is another exclusion criterion. Similarly, any potential participants requiring sedative drugs will not be included to avoid confounding factors related to brain activity that could alter the study’s findings.

Should a participant be diagnosed with a disease or dysfunction during the course of the study, they will be automatically excluded if the medical staff recommends their removal for health reasons. Additionally, the study protocol mandates that no external musical stimulation, including singing, be allowed for the enrolled participants during the admission period. Non-compliance with this guideline will result in the exclusion of the neonate from the study to maintain the integrity of the experimental conditions and data collection process.

#### Power calculation and sample size

In situations where an exact effect size is not derivable from prior research or pilot data – as in our case – utilizing the Minimum Effect Size of Interest (MESI) (Anvari & Lakens, 2021) is advantageous for guiding sample size estimation. This approach focuses on the smallest effect size that is considered practically or clinically significant within the specific context of a study, while it forms the foundation of the power analysis to ensure a study is calibrated to detect effects that are both meaningful and impactful.

For our study the MESI was established at Cohen’s *d*=0.3. This low determination was made in consultation with field experts and by reviewing clinical guidelines pertaining our research protocol, reflecting in result the minimal change in the primary outcome that would significantly impact patient care or treatment decisions in the specific context. More specifically, with our designated MESI for the musical intervention impacting premature infants’ brain activity, and by setting our alpha (Type I error) at 0.05 and power (Type II error) at 0.80, a power analysis with G*Power 3.1.9.7 suggested – at a High-Power Scenario – that 30 participants are needed to adequately power the study to detect this minimum effect size with desired statistical certainty (Table 1).

**Table 1.**
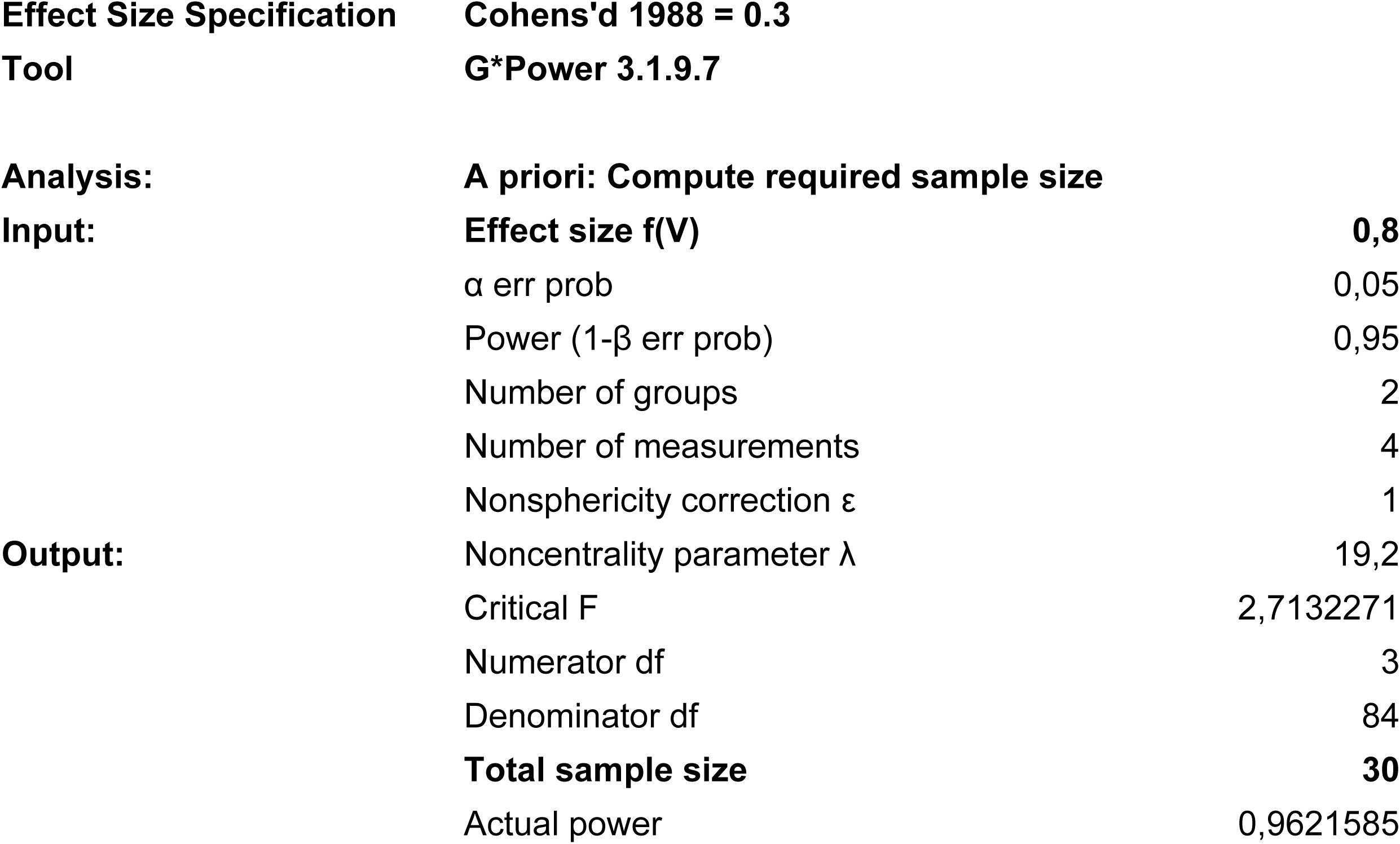
Power analysis table.

|  |  |  |
| --- | --- | --- |
| **Effect Size Specification** | **Cohens'd 1988 = 0.3** |  |
| **Tool** | **G\*Power 3.1.9.7** |  |
| **Analysis:** | **A priori: Compute required sample size** |  |
| **Input:** | **Effect size f(V)** | **0,8** |
| | $\alpha$ err prob | 0,05 |
| | Power (1-β err prob) | 0,95 |
|  | Number of groups | 2 |
|  | Number of measurements | 4 |
| | Nonsphericity correction $\epsilon$ | 1 |
| **Output:** | Noncentrality parameter $\lambda$ | 19,2 |
|  | Critical F | 2,7132271 |
|  | Numerator df | 3 |
|  | Denominator df | 84 |
|  | **Total sample size** | **30** |
|  | Actual power | 0,9621585 |

#### Intervention

Preterm newborns who meet the inclusion criteria, as explained above, will be exposed to a specific, in terms of content and exposure timing, musical intervention (explained in more detail below) four times in a calendar week – once per day. By conducting the intervention in the proposed way, we expect to observe and record the immediate and short-term neurophysiological effects of the musical stimuli on the infants’ brains, while ensuring that the data reflects the true impact of the intervention, free from external variables that might arise from a more prolonged or sporadic schedule. Furthermore, the specific exposure over this consistent period will facilitate a comprehensive understanding of any cumulative or evolving effects of the intervention across consecutive sessions, allowing for a detailed analysis of the EEG baselines recorded each one of the four times the intervention is delivered (also explained in more detail below). It should be also noted that the facilitators’ order of intervention delivery for each infant will be changed between participants to achieve better randomization and to avoid potential confounding variables due to a set order of intervention delivery.

As far as the musical intervention’s structure is concerned, this will take place in two main singing stages complemented by three distinctive blocks of silence, before, between and after each sung session as shown in the following list.

▪ * 180 seconds of silence (*Silence1; Baseline1*)
▪ <u>1st singing stage</u>: Signing a specific tone at 440 Hz, repeated *ad libitum* for a total of 60 seconds. (*NoteA*)
▪ * 180 seconds of silence (*Silence2; Baseline2*)
▪ <u>2nd singing stage</u>: Singing an original song suitable for the NICU environment in terms of structure, acoustic information composition, and volume level (dB), with a total duration of 90 seconds. (*Song*)
▪ * 180 seconds of silence (*Silence3; Baseline3*)

We decided to introduce these periods of silence before and after each singing stage to facilitate a return to the baseline of brain activity between the two singing stages, as well as to reduce the potential impact of any residual neural activity from the previous stage. This approach will enable us to achieve a more accurate assessment of the effects of each individual singing stage, and it will also allow for a clearer comparison of the developmental trajectory of the EEG baselines. By incorporating these controlled intervals of silence, we aim to isolate the neurophysiological effects specific to each musical intervention, thus providing a more precise understanding of how the infants’ brain activity evolves over the course of the intervention conditions as well as sessions.

In regard to the singing stages, as stated above, all facilitators will be asked to sing at the same distant location from the incubator, maintaining in this way consistent spatial sound parameters of stimulus delivery. For the first stage of the singing part (NoteA), each facilitator will be given a tuning fork (a forked acoustic component that produces sound upon impact and quickly silences) tuned to the musical note A (440 Hz) and asked to sing based on its produced sound the musical note A (440 Hz) as best as they can. All facilitators will need to repeat the specific note *ad libitum* over 60 seconds at this stage. The tuning fork has been chosen to be used as it is a safe acoustic accessory for accurately reproducing the tone the facilitator needs to hear as an acoustical template when inserting it into their ear, ensuring at the same time that the infant will not be able to hear it and be affected by it.

For the second singing stage (Song), each facilitator will need to sing a complex acoustic stimulus with organized sounds in the form of a lullaby, specifically created and composed for the study by a professional musician and music therapist (a member of the research team). This lullaby was aimed to have a unique structure and acoustic information composition adhering to the known scientific evidence related to infants’ sound and music perception capabilities (Háden et al., 2024; Trehub, 1987). The song lasts 90 seconds and features a soft melodic line, gentle rhythm, simple harmonies, and soft vocal timbre. Its tempo is expected to be approximately 60–75 beats per minute, following the rhythm of the seconds hand of the clock found on site as best as possible for consistency of delivery.

Finally, to ensure the best possible implementation of the intervention – particularly regarding the singing process – all parents of participants who meet the study’s inclusion criteria will receive 30 minutes of training before the actual intervention. This training will be conducted by a member of the research team who is a professional musician, singer, and music therapist. This pre-intervention session aims to equip parents with the necessary skills and understanding to enhance the intervention’s effectiveness, ensuring close consistency among the different facilitators. Additionally, the training session is designed to provide a calming influence on the parents, which can reduce stress and anxiety for both the infants and their caregivers during the delivery of the intervention. We believe that this pre-intervention training will empower parents as active participants in their child’s care, improving adherence and contributing to a more supportive and therapeutic environment throughout our study in the NICU.

#### Protocol Amendments

The study protocol reported here is the second version, following several adjustments made for both practical and scientific purposes. A process evaluation of the RCT was carried out in line with established medical research guidelines (French et al., 2020) and the findings informed a series of recommendations, which were incorporated into the current study design. A detailed account of the evaluation process as well as the changes made from the original protocol can be found in Dimitropoulos (2025).

#### Outcomes and Statistical analysis

To rigorously evaluate the effects of our musical intervention, we decided to employ a series of advanced statistical methods tailored to the specificities of the study design and data characteristics. As a result, the complete analysis was structured to address the research questions, focusing on the EEG-based outcome measures, with an emphasis on Spectral Power and Spectral Entropy, across the Delta frequency band.

Special attention was given to the Delta frequency band as it is widely recognized for its relevance in neonatal brain activity (van’t Westende et al., 2022; Bourel-Ponchel et al., 2021). The Delta frequency band, typically ranging from 0.5 to 4 Hz, is the slowest of the EEG frequency bands and is a critical marker of brain maturation. Higher absolute power in the delta frequency band has been associated with more favorable long-term neurodevelopmental outcomes, reflecting the progression of underlying neural processes during early development (van’t Westende et al., 2022). Given its prominence in this context, we believe that the Delta frequency band can be an essential marker for assessing the impact of sound and music on the neurophysiological state of our participants, providing in result critical insights into how these infants process auditory stimuli and the extent to which our intervention influences brain activity patterns.

More specifically to the design of our statistical analysis process and given the study’s repeated measures design and the nested data structure (with multiple observations per infant), a mixed-effects model was decided to be employed to analyze first the Delta Spectral Power, allowing for the inclusion of both fixed effects (conditions and facilitators) and random effects (variability across individual infants), while also meticulously fitting the model to account for the inter-individual variability after isolating the impact of the different intervention conditions and facilitator profiles. Specifically, to the Spectral Power approach, we decided to focus on this EEG feature as it can provide information about the intensity and prevalence of different types of neural oscillations (Gao, 2016) and it can help us better understand how the fluctuating premature infant brain activity changes over time. The thorough statistical analysis approach we propose here on the Delta Spectral Power is expected to generate fixed and random effects estimates, while to further elucidate the specific differences between condition/facilitator combinations, a Least Significant Difference (LSD) post-hoc test will be also conducted on the model’s outcome, aiming to identify which specific pairs of conditions or facilitators exhibit significant differences in brain activity, despite the increased risk of Type I errors due to multiple comparisons.

Parallel to the analysis of Delta Spectral Power, a similar mixed-effects model will be applied to the Delta Spectral Entropy data collected, following the same consideration for random and fixed effects, and aiming to explore how variations in spectral entropy, influenced by the different intervention profiles, reflect the neurophysiological responses of the infants. We decided to further study the EEG feature of Spectral Entropy in this context as it can offer a different dimension of analysis compared to Spectral Power and reflect in more detail the overall organization and complexity of the oscillations studied through the Delta Spectra Power approach. More specifically, Spectral Entropy is considered a measure of the complexity or irregularity of a signal’s frequency distribution (Kapucu et al., 2016), quantifying the degree of disorder or unpredictability within the power spectrum of a specific signal – in our case the Delta frequency band. A decrease in Spectral Entropy could indicate a transition to more synchronized and less complex neural activity, while conversely, higher Spectral Entropy might suggest more distributed and diverse neural activity, which could be linked to alertness or cognitive engagement.

Infusing this extra step of statistical analysis on our collected EEG data may be proved particularly valuable because it may reveal subtle changes in brain activity that may not be apparent through power analysis alone. As with the Spectral Power analysis, and subsequent to the mixed-effects model for Delta Spectral Entropy, a Least Significant Difference (LSD) post-hoc test will be also employed to investigate the differences between condition/facilitator combinations as in the previous approach.

#### Ethics

This study has been meticulously designed with a strong commitment to ethical standards, ensuring the protection and well-being of all participants. The research protocol has been thoroughly reviewed and approved by the ‘Panagiotis and Aglaia Kyriakou’ Hospital’s Scientific Council, under the Institutional Review Board (IRB) with protocol number 11333, 09th/04-05-2023 (Θ: 9). Additionally, the study has been registered on the clinical trials platform (www.clinicaltrials.gov) under the unique identifier NCT06398912, ensuring transparency and adherence to global research standards.

Participation in this study is entirely voluntary, and informed consent is a central component of the ethical process. Therefore, parents of identified premature infants who meet the inclusion criteria are invited to a detailed one-on-one meeting with the research team (NICU director), in order to receive comprehensive information, both orally and in writing, regarding the study’s objectives, methodologies, and procedures. This meeting covers all aspects of the intervention and data collection, including the non-invasive nature of the research tools, such as music and EEG, while also addresses any concerns related to selection, participation, withdrawal, confidentiality, and the study timeline, ensuring that parents are fully informed before making a decision.

During this meeting, the voluntary nature of participation is emphasized, with parents being assured that their decision to participate or withdraw at any point will not affect their relationship with the researchers or the medical staff at the Panagiotis & Aglaia Kyriakou Children’s Hospital. They are also informed that in the event of withdrawal, all data associated with the participant will be immediately deleted, ensuring no residual information remains.

Considering the sensitivity of the particular context, and to further protect the participants’ privacy and confidentiality, all collected data are de-identified and pseudonymized at the source (i.e., embeded function of the **nëo™** EEG monitoring unit during the extraction process), with each dataset assigned a unique identification number. Personal identifiers, such as names, are not linked to the data in any study analysis, report, or publication phase, while parents are informed of their right to access the data collected from their infants at any time. Once parents have received all relevant information and have had their questions answered, they are asked to provide written consent, indicating their agreement to participate in the study.

At this point, we should mention that despite its non-invasive nature, the study recognizes potential risks, such as the overstimulation of infants due to sound exposure and stress related to handling. To mitigate these risks, sound levels are closely monitored using a dB dosimeter – as already mentioned – and interventions are scheduled in this way so to ensure adequate rest periods for the infants. It has already been pointed above that the study is conducted in a specially designated, quiet room within the NICU to minimize stress, while moreover, the application of EEG electrodes, a key part of the study, is performed by specialists trained in neonatal care, further reducing the risk of discomfort or stress for the infants. In case of any unforeseen medical complications or adverse reactions, the study protocol includes provisions for the immediate cessation of the intervention and prompt medical evaluation, ensuring the safety of the participants.

#### Trial status

Since the ethical approval by the IRB and the Hospital Scientific Board in May 2023, 16 participants have been randomized and measured in our study – 11 in the experimental group and five in the control group. However, data from only five participants from the experimental group have been analyzed so far, comprising the basic participants workforce whereupon the EEG data pre-processing and processing pipelines, as well as the statistical analyses framework were first implemented. This small analysis cohort is consisted by two female and three male premature infants ranging between 28 and 32 weeks of gestation (i.e., specifically 199-227 days of gestation; SD = 12.42 days; Median (75^th^ Percentile) = 227 days). All five participants were measured in their 35th week PMA.

Prematurity in this dataset is underscored by the diverse maternal histories, including one mother who underwent in vitro fertilization (IVF) resulting in dichorionic diamniotic (DCDA) twin pregnancy (described as uneventful); a mother with an IgA nephropathy and a kidney transplantation, coupled with diabetes during pregnancy; one case involving increased resistance in the uterine arteries, raising concerns about potential intrauterine growth restriction (IUGR) and preeclampsia, and one last case experiencing a pregnancy complicated by IVF, gestational diabetes, polyhydramnios, and abnormal Doppler findings. Regarding the infants’ treatment, antibiotics and caffeine were commonly administered (typical neonatal care practices employed), with three cases receiving a standard combination of antibiotics and caffeine, while one case specifically involved ampicillin and gentamicin alongside caffeine. In one instance, no medication was reported, indicating a lack of need for such intervention.

In terms of the EEG analysis results for this dataset, the descriptive statistics for spectral power in the Delta frequency across different facilitators and conditions are detailed in table 2 below. The highest mean spectral power was observed during maternal interventions, particularly in the ‘NoteA’ condition (Mean = 1381.3 *μ*V^2^, SD = 1794.5 *μ*V^2^), emphasizing the notable influence of maternal presence in general.

**Table 2.**
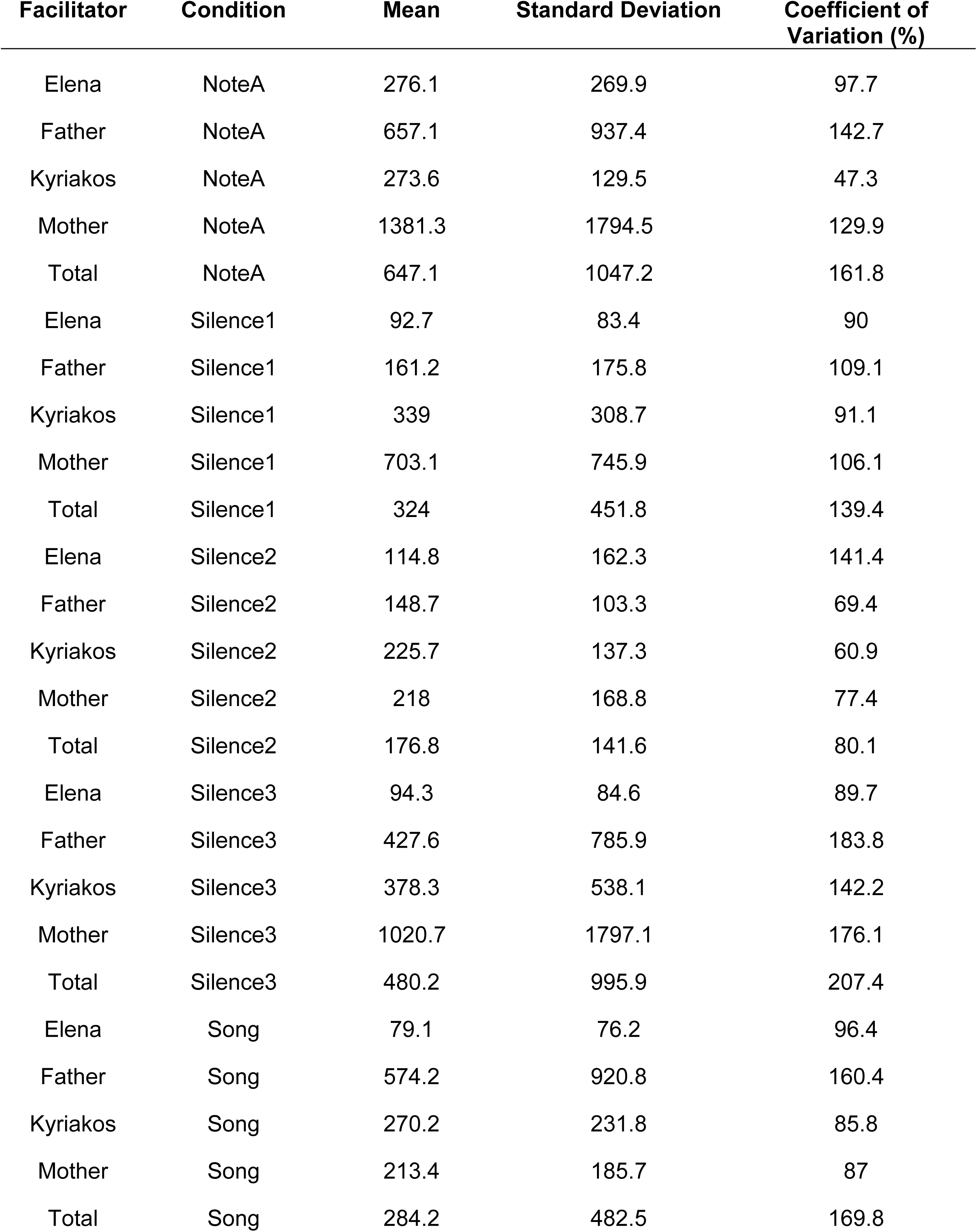
Descriptive statistics for Spectral Power on Delta frequency (in *μ*V^2^ for Mean and SD)

Following the descriptive statistics, and before running the mixed-effects model as already planned, an assumptions check was performed, creating a Q-Q plot to check for the normality of residuals and running the Breusch-Pagan test to check for any violation of homoscedasticity in the dataset. The Q-Q plot, as shown in figure 4, confirmed that the residuals followed an approximately normal distribution, while the Breusch-Pagan test for homoscedasticity resulted in a p-value of 0.066, suggesting on the one hand that the variance of residuals is consistent across different levels of the fixed effects, and indicating on the other hand that the assumption of homoscedasticity holds.

**Figure 4.**
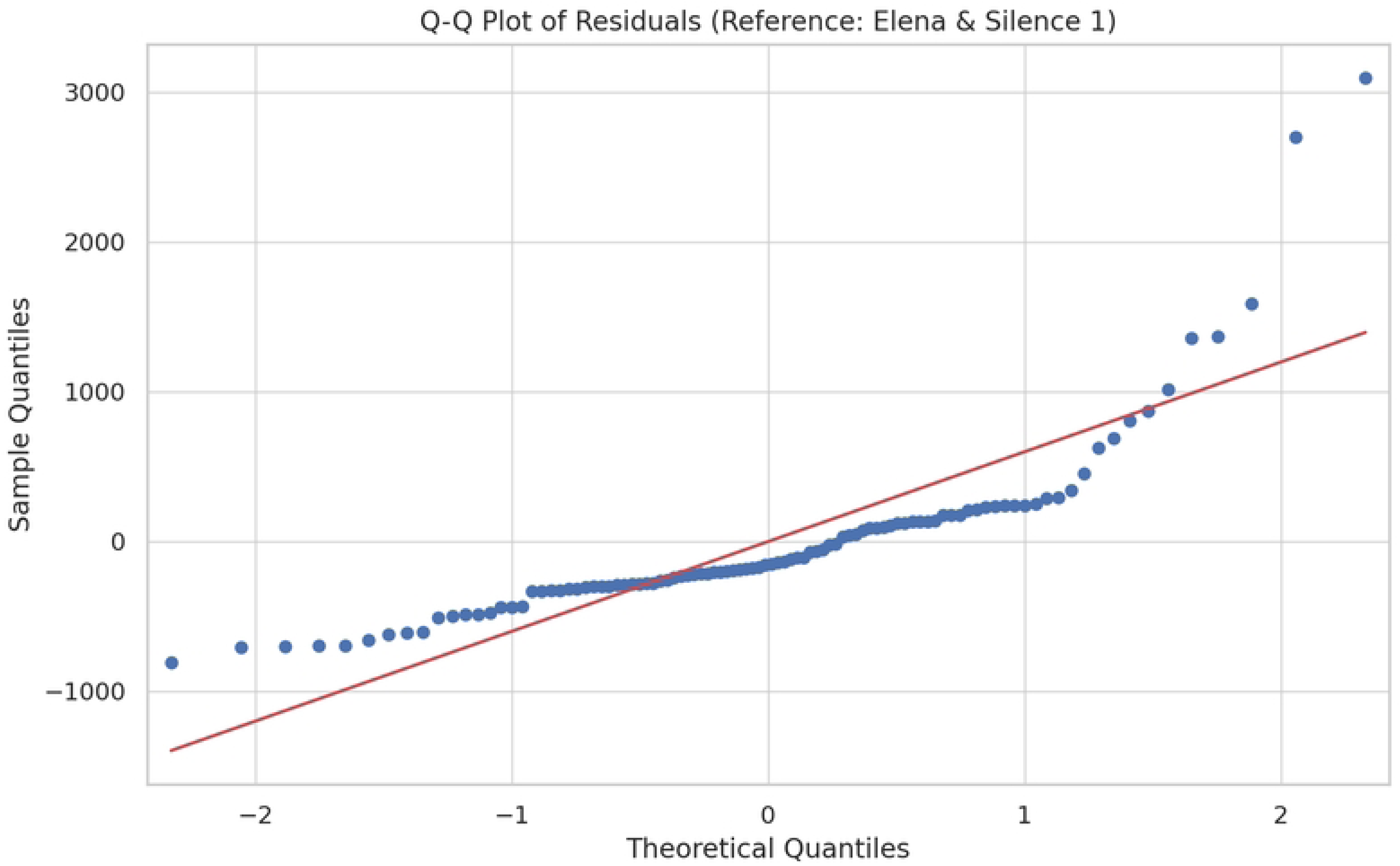
Q-Q Plot of Residuals with a reference point to the female music therapist for the facilitators and the ‘Silence1’ condition for the five intervention conditions

Considering the model assumptions to hold, we run first the mixed-effects model on the Spectral Power of the Delta frequency as decided, which included 100 observations across 5 groups, with each group comprising 20 observations at multiple time points. The model converged successfully with a scale parameter of 404,270.9768 and a log-likelihood of −739.3134. The coefficients for each condition and facilitator, along with their standard errors and p-values, are reported in full in table 3. The model revealed a significant effect of the ‘facilitator’ parameter on spectral power [ F(3,88) = 3.627, *p* = 0.016 ], while the ‘condition’ parameter did not show a statistically significant sole effect overall [F(4,88) = 1.669, *p* = 0.164)]. On the contrary, when we combined the effects across different conditions and facilitators (please see interaction plot in figure 5), the model revealed that although the Delta Spectral Power generally fluctuates, some interesting findings can be drawn based on this synthesis.

**Figure 5.**
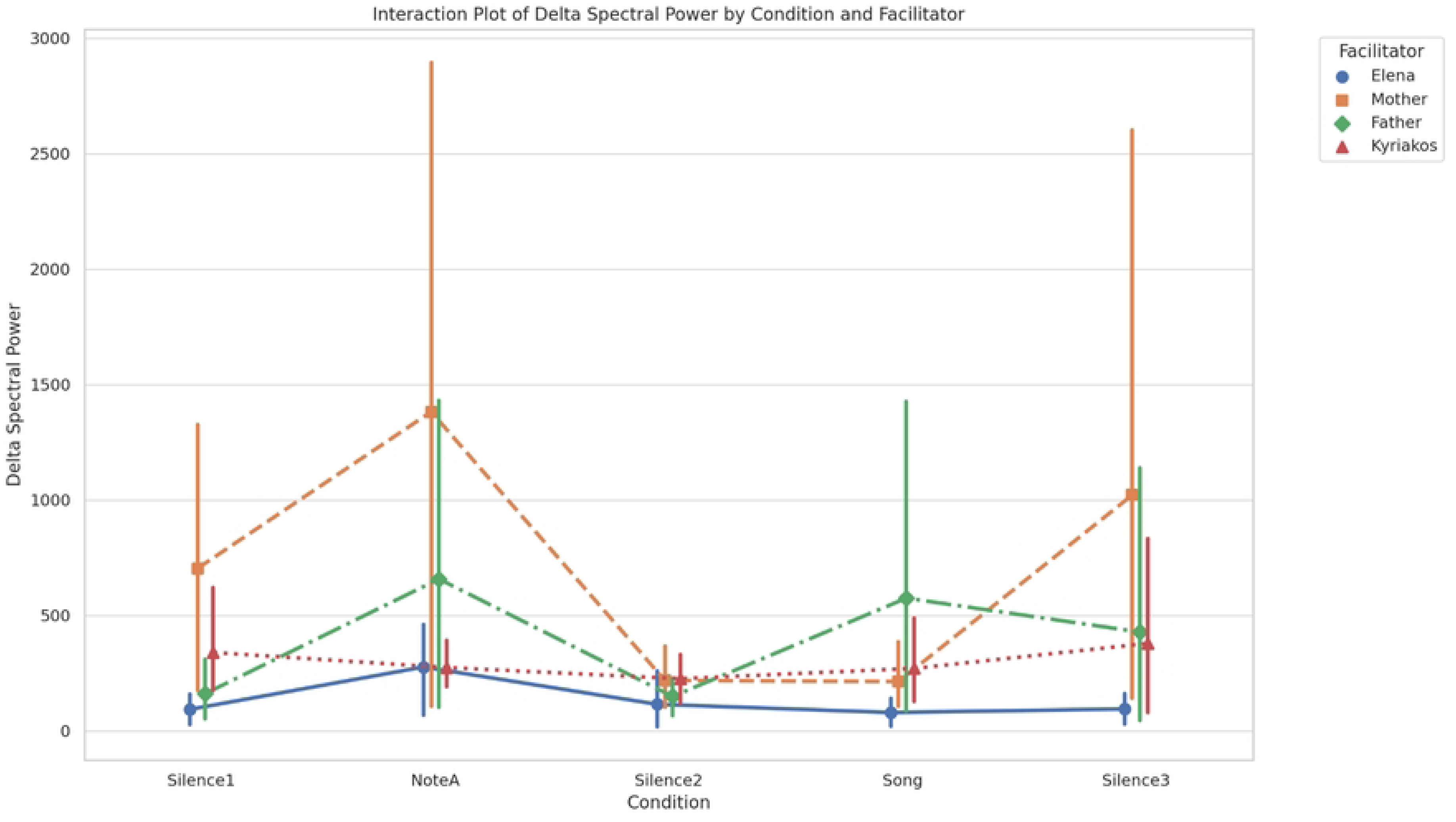
Combination of effects across different conditions and facilitators

**Table 3.** Coefficients for each condition and facilitator with SE and *p* values.

| Condition/Facilitator | Coefficient (Mean) | Standard Error | p-value |
| --- | --- | --- | --- |
| Elena | 131,442 | 181,968 | .148 |
| Father | 393,818 | 181,968 | .593 |
| Kyriakos | 297,398 | 181,968 | .025 |
| Mother | 707,362 | 181,968 | .002 |
| NoteA | 647,105 | 192,757 | .112 |
| Silence1 | 324,049 | 192,757 | .466 |
| Silence2 | 176,847 | 192,757 | .022 |
| Silence3 | 480,281 | 192,757 | .135 |
| Song | 284,243 | 192,757 | .075 |

More specifically, we realized that mostly during the ‘NoteA’ condition (as also observed in most of the other conditions), there was a significant increase in Delta Spectral Power for the ‘mother’ facilitator, while surprisingly, the ‘Song’ condition showcased the highest impact particularly for the ‘father’ facilitator. As far as the ‘female music therapist (Elena)’ facilitator is concerned, the most stable and lowest Delta Spectral Power across all conditions was observed, indicating a consistent but minimal impact, while the ‘male music therapist (Kyriakos)’ facilitator showed moderate variability with minor peaks during the ‘NoteA’ and ‘Song’ conditions. Please note here, that especially for the ‘Song’ condition, the ‘male music therapist (Kyriakos)’ facilitator ranked second in impact after the ‘father’ facilitator. To further clarify these specific differences, post-hoc comparisons using the Least Significant Difference (LSD) test were conducted as previously decided, revealing that Delta brain activity was significantly higher with the ‘Mother’ facilitator compared to ‘Elena’ (M = 575.92, *p* = 0.002), ‘Kyriakos’ (M = 409.96, *p* = 0.025), and ‘Father’ (M = 313.55, *p* = 0.085) – although for this latter coming quite close to being significant. Additionally, the ‘NoteA’ condition elicited significantly higher brain activity than the ‘Silence2’ condition (M = 470.26, *p* = 0.022) and showed close trends towards significance when compared to the ‘Silence1’ (M = 323.06, *p* = 0.112) and ‘Song’ conditions (M = 362.86, *p* = 0.075).

Finally, as already mentioned, in addition to the analysis of Delta spectral power, we also explored the spectral entropy within the Delta frequency band to assess the complexity and predictability of the EEG signals under different facilitators and conditions. The mixed-effects model analysis for spectral entropy revealed once more a significant effect of the ‘facilitator’ parameter on entropy values [F(3, 88) = 2.744, *p* = 0.048], while the spectral entropy across the ‘conditions’ parameter did not exhibit a significant overall effect [F(4, 88) = 0.380, *p* = 0.822]. Especially for the former result, underscoring that the facilitator’s role notably influences the variability and regularity of the EEG signals, we realized that, specifically, the ‘father’ facilitator was associated with the highest mean entropy (M = 0.684, SE = 0.133), while the ‘mother’ facilitator showed the lowest mean entropy (M = 0.558, SE = 0.133) (please see also figure 6); although pairwise comparisons did not reach statistical significance (in all cases except the close to significance trend of the ‘father’ vs ‘mother’ comparison) after Bonferroni correction (please see table 4)

**Figure 6.**
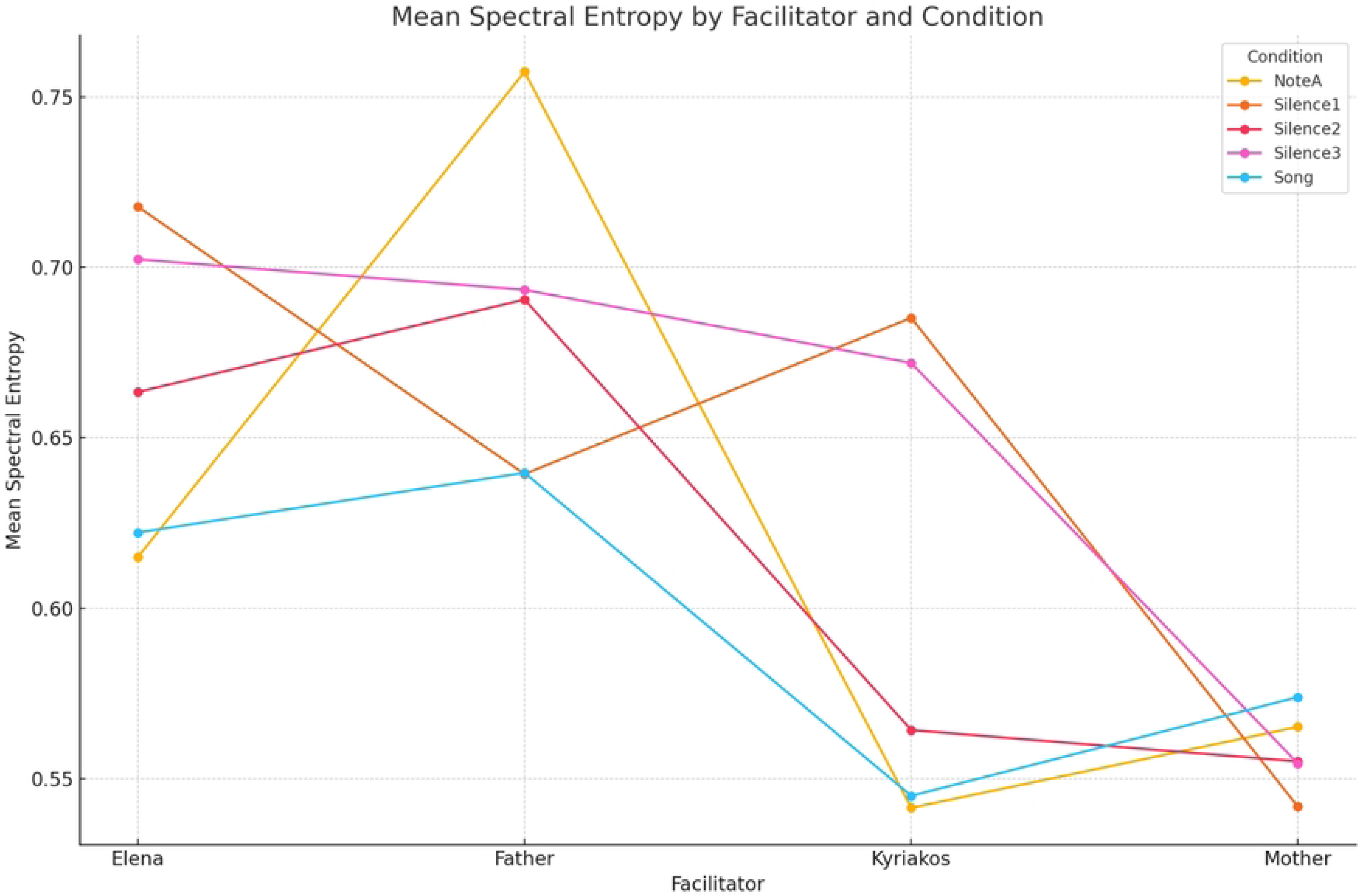
Mean spectral entropy within the Delta band, showing a significant effect of the ‘facilitator’. The ‘father’ shows the highest mean entropy and the ‘mother’ the lowest.

**Table 4.** Pair-wise comparison of the ‘facilitator’ condition after Bonferroni correction.

| Comparison | Mean Difference | Standard Error (SE) | p-value |
| --- | --- | --- | --- |
| Elena vs. Father | -0.02 | 0.049 | 1 |
| Elena vs. Kyriakos | 0.063 | 0.049 | 1 |
| Elena vs. Mother | 0.106 | 0.049 | 0.207 |
| Father vs. Kyriakos | 0.083 | 0.049 | 0.589 |
| Father vs. Mother | 0.126 | 0.049 | 0.075 |
| Kyriakos vs. Mother | 0.043 | 0.049 | 1 |

## Discussion

Although there is emerging evidence that musical interventions can positively influence the neurodevelopment of premature infants, existing studies often suffer from limited control over confounding variables, and a lack of rigorous neuroimaging-based evidence to support the precise neural mechanisms underlying these effects. Against this backdrop, the aim of the present study is to fill these gaps by providing robust, RCT-based data on the effects of a structured singing intervention, specifically focusing on the fundamental frequency (F0) of the voice and kinship of the music facilitators parameters, on the brain activity of premature infants in a Neonatal Intensive Care Unit (NICU) setting. Building upon this foundation, some of the key strengths of the protocol presented here include: (1) the use of an RCT design with an adequately powered sample size to detect statistically significant differences in electroencephalographic (EEG) activity across different conditions and facilitators; (2) the employment of an advanced neuroimaging technique to precisely monitor and study neural correlates of the intervention; and (3) the consideration of a variety of caregivers – in terms of kinship and voice register – to deeper explore the specific impact of singing on premature infant brain development.

More specifically, we must emphasize, first of all, that this study protocol was meticulously designed as a randomized controlled trial (RCT) – including, therefore, both an experimental and a control group in its design – in order to provide a solid foundation for causal inferences regarding the effects of musical interventions on the brain activity of premature infants, allowing for a more nuanced understanding of any embedded confounders. Although our work does not yet report on the statistical comparisons between these groups, we believe that the approach we take is crucial for discerning any differences in the measurement process and the application of the specific stimuli, yielding more profound insights into the efficacy of the intervention. Moreover, with the integration of neuroimaging and more specifically the custom-made and adapted EEG (pre)processing pipeline utilized, the effects of our musical intervention on premature infants can be better studied and analyzed in more detail, thus playing a pivotal role in understanding the fine nuances of implementing the tools employed in the specific context. As discussed in previous work (Papatzikis, 2024), neuroimaging-guided interventions should be more extensively introduced in the NICU because they can provide real-time insights into the brain’s response to external stimuli, capturing in essence dynamic changes that might not be observable through behavioral measures alone. In this regard, our protocol approach not only enhances the precision of our study but can also contribute to the development of more tailored applications of this sort in the near future, optimizing other similar music-based interventions to suit the unique neurodevelopmental needs of each infant in the NICU.

Finally, it is important to point out that the study design we decided to implement includes the chosen variety of caregivers to allow us to provide a more comprehensive insight into the role of emotional and social factors evident and embedded in the treatment process of premature infants in real-word situations and environments. For that matter, on the one hand, the inclusion of both parents in the protocol ensures that the study captures potential sex-specific effects, which could be related to differences in voice register or emotional bonding, while, on the other hand, incorporating professional music therapists as facilitators offers a standardized comparison to the former set of caregivers, helping to differentiate the effects of kinship as well as musical and clinical expertise related to the implementation of music-based interventions in this sensitive context. This diversity in caregivers could not only enrich the study’s findings but may also enhance the generalizability of its results, ultimately leading to more comprehensive recommendations for NICU music-based interventions.

However, despite the strengths of our study, certain challenges and limitations must also be acknowledged. First, the heterogeneity of the study population, which may potentially include infants with varying degrees of prematurity – as defined by their gestational age – and differing health profiles, may introduce significant variability and complicate the interpretation of the results, as the infants’ responses to the musical interventions may be influenced by factors beyond the scope of the study. For instance, the inclusion of participants with diverse medical histories, while increasing the generalizability of the findings, also necessitates careful consideration in data analysis and further clinical implementation to account for these differences. Furthermore, external factors, such as the NICU environment, could also influence the study’s conduct and the quality of the data collected. The acoustic environment for once, despite being controlled within the study’s dedicated room, may still introduce variability that could affect the infants’ responses, while the fragile emotional and psychological state of the parents, who are also facilitators in the study, could impact the delivery and reception of the musical intervention. These factors, although managed to the greatest extent possible, remain potential sources of variability that could influence the outcomes.

Despite these challenges, the potential impact of this study remains highly significant. Its findings, for once, could be easily integrated into existing neonatal care practices, offering healthcare providers a scientifically grounded method for enhancing the care of premature infants. Furthermore, the broader impact of this study could be substantial as the insights gained from the differential effects of maternal, paternal, and therapist-facilitated singing on neonatal brain activity could inform wider public health strategies aimed at improving neurodevelopmental outcomes for premature infants. Finally, this research may also encourage the expansion of music-based interventions in neonatal care units worldwide, particularly in regions where such practices are not yet widely implemented.

## Conclusion

As this study is still in progress, it is essential to recognize that the findings presented here are preliminary, based on an initial analysis cohort. Therefore, the continuation and completion of the study will be critical for validating these early results and for providing a more comprehensive understanding of the effects of the specific music-based intervention on neonatal brain development. That said, several areas for further investigation have emerged from the process of structuring, fine-tuning, and implementing our study protocol, suggesting for future research to explore the long-term effects of this and other similar interventions, particularly in terms of cognitive and behavioral outcomes as the infants develop. Additionally, more research is needed to understand the specific mechanisms through which other dynamic soundscapes and modes of musical delivery may influence the infants’ brain activity, leading to more targeted and effective interventions. For all the above, it is our hope that the detailed report of our study protocol and its preliminary results provided here, may help future researchers better accommodate their investigation needs and study hypotheses in the emerging field of music-based interventions in the NICU with the help of neuroimaging tools and particularly electroencephalography, leading to new horizons of applicable treatment in this very sensitive yet crucial environment for human development.

## Data Availability

Data cannot be shared publicly because of the infant population included in the measurement process. Data are available from the Director of the NICU or the Lead Author of the manuscript for researchers who meet the criteria for access to confidential data.

## Notes

### Competing Interest Statement

The authors have declared no competing interest.

### Clinical Trial

clinincaltrials.gov unique identifier: NCT06398912

### Funding Statement

The author(s) received no specific funding for this work.

### Author Declarations

The research protocol has been thoroughly reviewed and approved by the ‘Panagiotis and Aglaia Kyriakou’ Hospital's Scientific Council, under the Institutional Review Board (IRB) with protocol number 11333, 09th/04-05-2023 (Θ: 9). Informed consent to participate in this study was obtained both orally and in writing.

